# Dynamics of T-cell responses following COVID-19 mRNA vaccination and breakthrough infection in older adults

**DOI:** 10.1101/2023.07.14.23292660

**Authors:** Sneha Datwani, Rebecca Kalikawe, Francis Mwimanzi, Sarah Speckmaier, Richard Liang, Yurou Sang, Rachel Waterworth, Fatima Yaseen, Hope R. Lapointe, Evan Barad, Mari L. DeMarco, Daniel T. Holmes, Janet Simons, Julio S.G. Montaner, Marc G. Romney, Zabrina L. Brumme, Mark A. Brockman

## Abstract

**Introduction:** While older adults generally mount weaker antibody responses to a primary COVID-19 vaccine series, T-cell responses remain less well characterized in this population. We compared SARS-CoV-2 spike-specific T-cell responses after two- and three-dose COVID-19 mRNA vaccination and subsequent breakthrough infection in older and younger adults.

**Methods:** We quantified CD4+ and CD8+ T-cells reactive to overlapping peptides spanning the ancestral SARS-CoV-2 spike protein in 40 older adults (median age 79) and 50 younger health care workers (median age 39), all COVID-19 naive, using an activation induced marker assay. T-cell responses were further assessed in 24 participants, including 8 older adults, who subsequently experienced their first SARS-CoV-2 breakthrough infection.

**Results:** A third COVID-19 mRNA vaccine dose significantly boosted spike-specific CD4+ and CD8+ T-cell frequencies to above two-dose levels in older and younger adults. T-cell frequencies did not significantly differ between older and younger adults after either dose. Multivariable analyses adjusting for sociodemographic, health and vaccine-related variables confirmed that older age was not associated with impaired cellular responses. Instead, the strongest predictors of CD4+ and CD8+ T-cell frequencies post-third-dose were their corresponding post-second-dose frequencies. Breakthrough infection significantly increased both CD4+ and CD8+ T cell frequencies, to comparable levels in older and younger adults. Exploratory analyses revealed an association between HLA-A*02:03 and higher post-vaccination CD8+ T-cell frequencies, which may be attributable to numerous strong-binding HLA-A*02:03-specific CD8+ T-cell epitopes in spike.

**Conclusion:** Older adults mount robust T-cell responses to two- and three-dose COVID-19 mRNA vaccination, which are further boosted following breakthrough infection.

## INTRODUCTION

In many jurisdictions, older adults were prioritized to receive COVID-19 vaccines and boosters due to their increased risk of severe outcomes following SARS-CoV-2 infection^1–3^. While vaccination has been highly effective at preventing severe disease in this group^4^, vaccine responses in older adults can nevertheless be blunted by age-related immune impairments, or elevated frequencies of chronic health conditions that can dampen adaptive immune responses^5–9^. Indeed, after COVID-19 vaccines were rolled out globally, observational studies revealed that older adults generally mounted weaker binding and neutralizing antibody to the primary vaccine series^10–15^, leading to widespread recommendations that this group receive third vaccine doses and regular booster vaccinations^16, 17^. Comparably fewer studies however have investigated cellular immune responses to COVID-19 vaccination − namely, CD4+ helper T cells that play a central role in the generation of antigen-specific B cells and antibody responses, and CD8+ cytotoxic T cells that recognize and eliminate virus-infected cells^18^ − in older compared to younger adults. While mRNA vaccines can induce strong T cell responses^19, 20^, evidence suggests that the frequency of spike-specific CD4+ T cells following COVID-19 vaccination may be lower in older adults^11, 13, 21^. An improved understanding of age-associated differences in T cell responses to COVID-19 mRNA vaccines will help to inform future efforts to enhance protective immunity in older adults. Here, we investigated the dynamics of spike-specific CD4+ and CD8+ T cell responses elicited after two and three doses of COVID-19 mRNA vaccine in a cohort of 40 older adults and 50 younger healthcare workers who remained naive to SARS-CoV-2 during this time. We additionally investigated spike-specific CD4+ and CD8+ T cell responses in a subset of 24 individuals, including 8 older adults, who subsequently experienced their first SARS-CoV-2 breakthrough infection between one and six months after receiving three vaccine doses. Finally, we explored associations between HLA class I allele carriage and the magnitude of spike-specific CD8+ T cell frequencies after two and three COVID-19 mRNA vaccine doses.

## METHODS

### Participants

Our cohort, based in British Columbia (BC) Canada, has been described previously ^22^. Here, we studied a randomly selected subset of 50 healthcare workers (HCW) and 40 older adults (OA, age >65 years) who remained COVID-19 naive until at least one month after their third COVID-19 mRNA vaccine dose (**Table 1**).

**Table 1:**
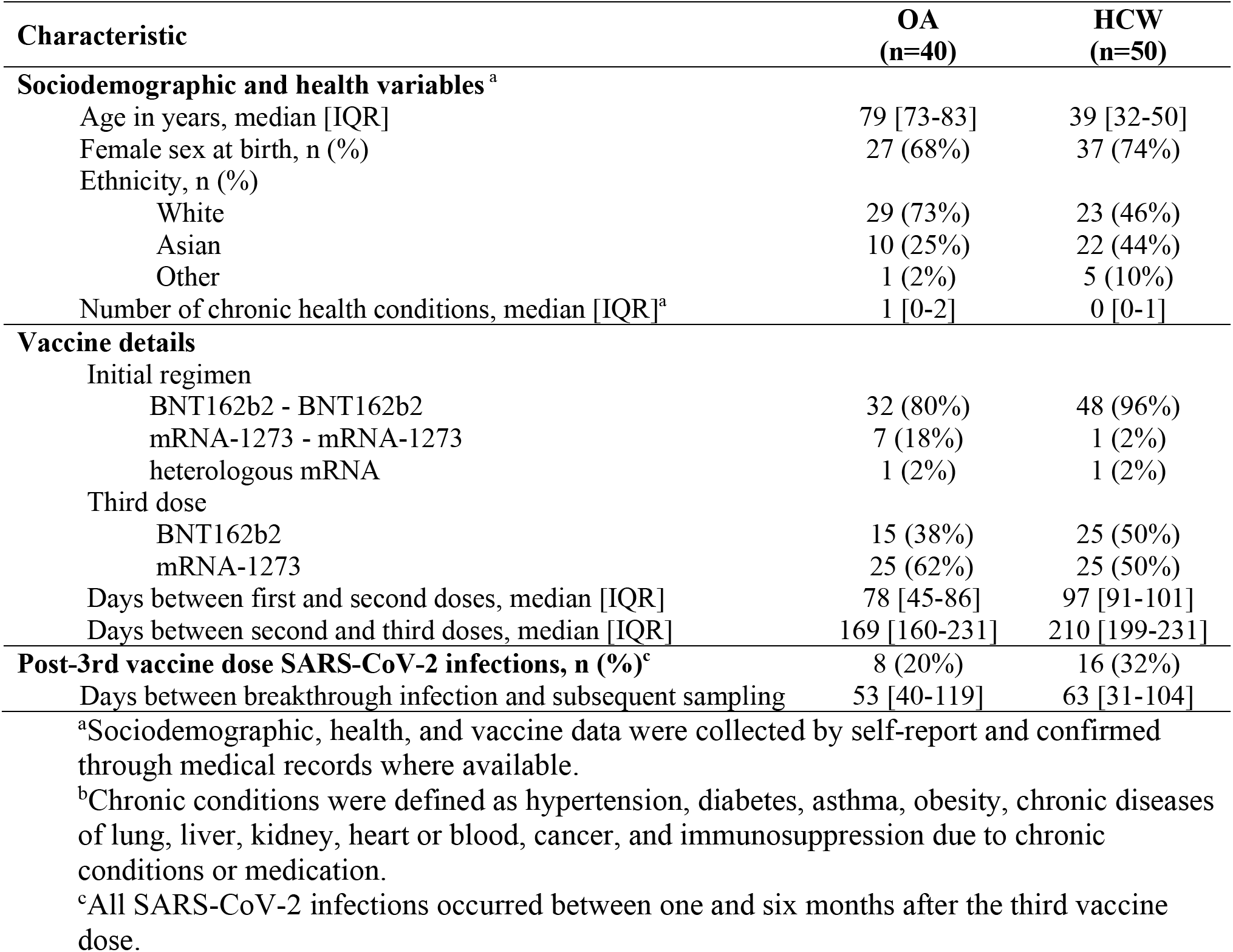
Participant characteristics.

### Ethics approval

Written informed consent was obtained from all participants or their authorized decision makers. This study was approved by the University of British Columbia/Providence Health Care and Simon Fraser University Research Ethics Boards.

### Antibody assays

We had previously quantified IgG-binding antibodies in serum against the ancestral SARS-CoV-2 spike receptor binding domain (RBD) using the V-plex SARS-CoV-2 (IgG) ELISA kit (Panel 22; Meso Scale Diagnostics)^14, 15, 22^. Serum was diluted 1:10000 and reported in World Health Organization (WHO) International Standard Binding Antibody Units (BAU)/mL using the manufacturer-supplied conversions. SARS-CoV-2 infections were detected by the development of serum antibodies against Nucleocapsid (N) using the Elecsys Anti-SARS-CoV-2 assay (Roche Diagnostics), combined with diagnostic (PCR- and/or rapid-antigen-test-based) information where available.

### T-cell assays

Cryopreserved peripheral blood mononuclear cells (PBMCs) were thawed and diluted in TexMACS media (Miltenyi Biotec, Cat#130-097-196). PBMCs were stimulated at 1 x 10^6^ cells per well for 24 hours with peptide pools spanning the SARS-CoV-2 ancestral spike protein (15-mers, overlapping by 11 amino acids) (Miltenyi Biotec, Cat#130-127-953) in duplicate in a 96-well U-bottom plate. PBMC were incubated with DMSO only (no peptide) as a negative control and 2 µg/ml Cytostim reagent (Miltenyi Biotec, Cat#130-092-172) as a positive control. Following stimulation, cells were labeled with CD8-APC/Cyanine7 (Biolegend, Cat#301016), CD4-FITC (Biolegend, Cat#300538), CD137-APC (Biolegend, Cat#309810), CD69-PE (BD, Cat#555531), OX40-PE-Cy7 (Biolegend, Cat#350012), CD3-PerCP/Cyanine5.5 (Biolegend, Cat#317336) CD14-V500 (BD, Cat#561391), CD19-V500 (BD, Cat#561121) and 7-AAD Viability Staining Solution (Biolegend, Cat#420404). Data were acquired on a Beckman Coulter Cytoflex flow cytometer, with a minimum of 10,000 CD3+ T cells assayed per participant. After identifying CD3+CD4+ and CD3+CD8+ T cell subsets, the percentage of stimulated cells was determined based on upregulation of activation markers, using CD137 and OX40 for CD4+ T cells and CD137 and CD69 for CD8+ T cells (see gating strategy in **Figure S1**). Data were analyzed in FlowJo version 10.8.1.

### HLA class I genotyping

Genomic DNA was isolated from 50 μl whole blood using the NucliSENS EasyMag system (BioMerieux). HLA class I genotyping was performed by locus-specific Polymerase Chain Reaction (PCR) amplification of the region spanning exons 2 and 3 of the HLA-A, B and -C loci, as reported previously^23^. Amplicons were bi-directionally sequenced on an ABI 3730xl automated Sanger DNA sequencer using BigDye (v3.1) chemistry (Applied Biosystems). Chromatograms were analyzed using the semiautomatic base-calling software RECall^24^, with the resulting bulk sequences interpreted to subtype-level resolution using in-house software.

### Epitope prediction

Peptides derived from ancestral spike protein (GenBank: NC_045512.2) that are likely to bind to HLA-A*02:01 and/or A*02:03 were predicted using NetMHCpan-4.1 (https://services.healthtech.dtu.dk/services/NetMHCpan-4.1; ^25^). Potential 8-, 9-, and 10-amino acid epitopes were determined for both alleles using default thresholds for strong binders (% Rank <0.5) and weak binders (% Rank 0.5-2).

### Statistical analyses

Continuous variables were compared using the Mann-Whitney U-test (for unpaired data) or Wilcoxon test (for paired measures). Relationships between continuous variables were assessed using Spearman’s correlation. Zero-inflated beta regressions were used to investigate the relationship between age and vaccine-induced T-cell responses using a confounder model that adjusted for variables that could influence these responses, or that differed in prevalence between groups. These regressions model the response variable as a beta-distributed random variable whose mean is given by a linear combination of the predictor variables (after a logit transformation). Beta distributions are bounded below and above by 0 and 100%, making this a standard choice of regression for frequency data. A beta distribution however does not admit values of 0 or 100. As our data included some non-responders (*i.e.* 0 values) we used a zero-inflated beta distribution, which allows for zeros in the data. For analyses performed after two-dose vaccination, included variables were: age (per year), sex at birth (female as reference), ethnicity (non-white as reference), number of chronic conditions (per additional), mRNA-1273-containing initial vaccine regimen (dual BNT162b2 vaccination as reference) and the interval between first and second doses (per day). Analyses performed after three-dose vaccination also included the third COVID-19 mRNA dose brand (BNT162b2 as reference), the interval between second and third doses (per day), and the % spike-specific T-cells after two doses (per percent increase). All tests were two-tailed, with p<0.05 considered statistically significant. For the analyses of the relationships between HLA class I allele carriage, multiple comparisons were addressed using a q-value (false discovery rate) approach^26^, with associations p<0.05 and q<0.2 considered statistically significant. Analyses were conducted using Prism v9.2.0 (GraphPad) and in R.

## RESULTS

### Participant characteristics

Characteristics of the 40 older adults (OA) and 50 younger health care workers (HCW), all of whom remained COVID-19 naive until at least one month after their third vaccine dose, are shown in **Table 1**. OA and HCW were a median of 79 and 39 years old, respectively (overall range 24-93 years old), and predominantly female. OA were predominantly of white ethnicity (73%, compared to 46% of HCW) and had more chronic health conditions (median of 1, interquartile range [IQR] 0-2 in OA versus 0 [IQR 0-1] in HCW). Most participants (80% of OA and 96% of HCW) initially received two doses of BNT162b2; the remainder received two doses of mRNA-1273 or a heterologous mRNA vaccine regimen. Second doses were administered a median of ∼3 months after the first. Third vaccine doses were predominantly mRNA-1273 (62% of OA and 50% of HCW), which were administered in an age-dependent manner per local guidelines: OA were eligible for 100 mcg whereas HCW were eligible for 50 mcg (all BNT162b2 doses were 30 mcg). Third doses were administered a median of 6 months after the second. A total of 20% of OA and 32% of HCW experienced their first SARS-CoV-2 infection between one and six months post-third dose, where these were likely Omicron BA.1 or BA.2 infections based on local molecular epidemiology trends^27^. The post-infection follow-up visit occurred 53 (IQR 40-119) days later for OA and 63 (IQR 31-104) days later for HCW.

### CD4+ and CD8+ T-cell responses following two and three COVID-19 vaccine doses

Prior to vaccination, the percentage of spike-specific CD4+ and CD8+ T cells, measured in a subset of 10 OA and 10 HCW, was negligible (**Figures 1A, 1B**). Following two vaccine doses, the percentage of spike-specific CD4+ T cells increased significantly from pre-vaccine levels (paired measures p=0.002 for both OA and HCW), reaching a median 0.25% (IQR 0.14-0.57%) in OA and a median 0.42% (IQR 0.14-0.66%) in HCW, a difference that was not statistically significant between groups (p=0.28; **Figure 1A**). A third vaccine dose further boosted the percentage of spike-specific CD4+ T-cells (paired measures p<0.0001 for both OA and HCW), reaching a median 0.62% (IQR 0.27-0.91) in OA compared to a median 0.77% (IQR 0.45-1.08) in HCW, a difference that was not statistically significant between groups (p=0.07) (**Figure 1A**).

**Figure 1.**
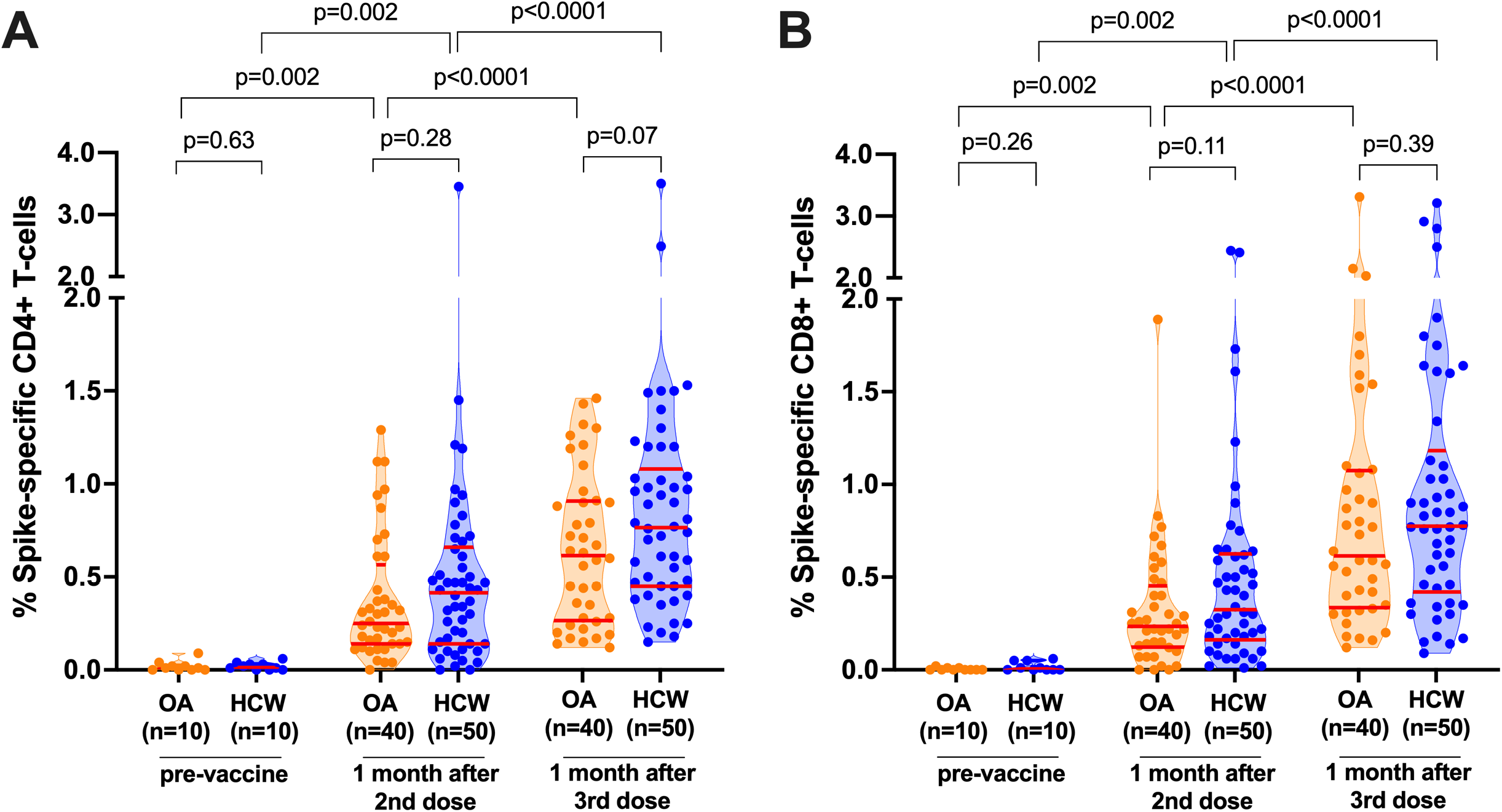
SARS-CoV-2 spike-specific T-cell frequencies before and after COVID-19 mRNA vaccination. *Panel A:* Spike-specific CD4+ T-cell frequencies before and after two and three-dose COVID-19 vaccination. Older Adults (OA) are in orange; younger Health Care Workers (HCW) are in blue. All participants are COVID-19 naive. Red bars indicate median and IQR. The Mann-Whitney U-test was used for between-group comparisons and the Wilcoxon matched pairs test was used for longitudinal paired comparisons. P-values are not corrected for multiple comparisons. *Panel B:* Same as panel A, but for spike-specific CD8+ T-cell frequencies.

CD8+ T cell responses also increased significantly from pre-vaccine levels following two vaccine doses (paired measures p=0.002 in both groups), reaching a median 0.24% (IQR 0.13-0.45%) in OA and a median 0.33% (IQR 0.16-0.63%) in HCW, a difference that was not statistically significant between groups (p=0.11; **Figure 1B**). A third vaccine dose further boosted the percentage of spike-specific CD8+ T-cells (paired measures p<0.0001 for both groups) to a median 0.62% (IQR 0.34-1.08%) in OA and a median 0.78% (IQR 0.42-1.18%) in HCW, a difference that was not statistically significant between groups (p=0.39; **Figure 1B**).

We observed no correlation between age and the percentage of spike-specific CD4+ or CD8+ T-cells after either the second or third vaccine dose, when age was assessed as a continuous variable (Spearman’s ρ ranged from = −0.14 to −0.03; all p>0.20; not shown). Moreover, we confirmed that age remained not significantly associated with vaccine-induced T-cell responses after adjusting for relevant sociodemographic, health and vaccine-related variables (**Tables S1 and S2**). Rather, the strongest predictor of the % of spike-specific CD4+ and CD8+ T-cells following three vaccine doses was the corresponding % of spike-specific T-cells following two doses (**Table S2**). For example, the zero-inflated beta regression estimates for the impact of each 1% increment in post-second dose T-cell frequencies on post-third dose frequencies was 0.69 for CD4+ T-cells (p=2×10^-16^) and 0.77 for CD8+ T-cells (p=1×10^-13^) (**Table S2**). To interpret these estimates, which do not translate the effects of the predictors linearly to the predicted mean: if the value of the linear predictor was x for a given set of values of the predictors, and the % of spike-specific CD4+ T-cells after two vaccine doses was increased by 1%, the resulting value of the linear predictor would now be x + 0.69, which translates to a non-linear increase in the regressed mean from 100 * logit(x) to 100 * logit(x + 0.69), where the factor of 100 converts the proportions to percentages. Enhanced CD4+ T-cell frequencies after three vaccine doses were also associated with male sex, having received mRNA-1273 as a third vaccine dose, and − somewhat surprisingly − a higher number of health conditions, though these correlates were all far weaker than the post-second-dose T cell responses (estimates 0.13-0.23; p=0.03 to 0.006; **Table S2**). We hypothesize that association between more health conditions and better responses is because individuals with such conditions benefited particularly from a third dose, after adjusting for post-second-dose responses. White ethnicity was also weakly associated with lower CD4+ T-cell frequencies after two vaccine doses (estimate −0.38; p=0.03; **Table S1**).

### Correlations between vaccine-induced humoral and T-cell responses

The percentage of spike-specific CD4+ and CD8+ T cells correlated strongly with one another after the second vaccine dose (Spearman’s ρ=0.60; p<0.0001) as well as after the third dose (Spearman’s ρ=0.59; p<0.0001; **Figure 2A**). No correlation however was observed between the magnitude of the COVID-19 vaccine antibody response, in terms of serum anti-spike Receptor Binding Domain (RBD) IgG concentrations after either the second or third vaccine dose, and the percentage of spike-specific CD4+ T-cells at those times (Spearman’s ρ = −0.09, p=0.41 for post-second dose; ρ = −0.11, p=0.32 for post-third dose; **Figure 2B**).

**Figure 2.**
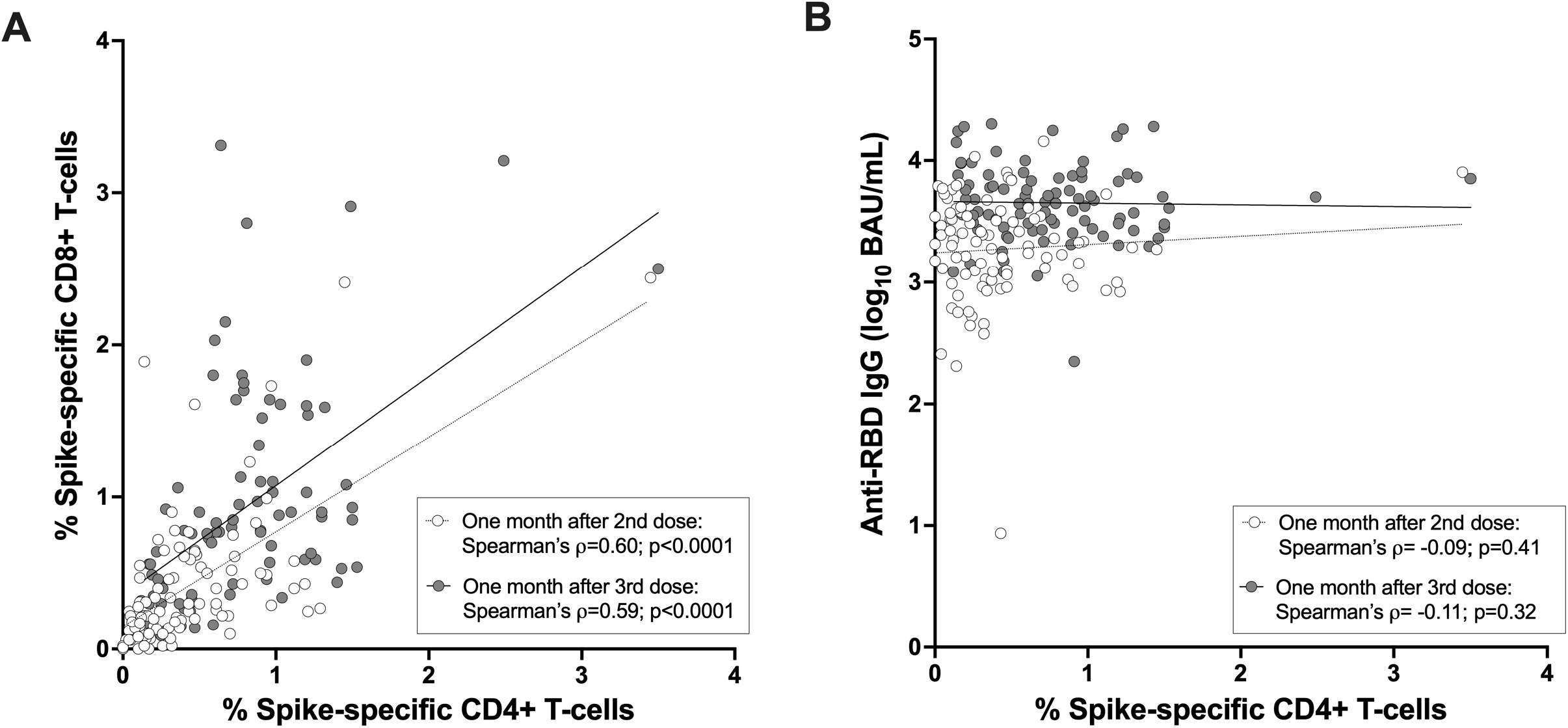
Correlations between cellular and humoral immune measures after two and three-dose COVID-19 mRNA vaccination. *Panel A:* Correlation between spike-specific CD4+ and CD8+ T-cell frequencies one month after the second dose (open circles) and one month after the third dose (closed circles) in the combined cohort. *Panel B*: Correlation between spike-specific CD4+ T-cell frequencies and spike-specific IgG binding antibodies, measured in one month after the second dose (open circles) and one month after the third dose (closed circles) in the combined cohort. All participants are COVID-19 naive.

### Correlations between HLA class I genotypes and CD8+ T-cell responses

Participants expressed a total of 29 different HLA-A, 42 HLA-B and 23 HLA-C alleles, defined at the subtype level (**Figure S2).** Exploratory analyses that included all HLA alleles observed in a minimum of 5 participants revealed that, following three vaccine doses, expression of HLA-A*02:03 was associated with higher spike-specific CD8+ T cell response frequencies after correcting for multiple comparisons (p=0.01; q=0.08), while expression of B*39:01, B*44:03, or C*07:01 was associated with lower spike-specific CD8+ T cell response frequencies (all p<0.03, q<0.13) (**Table S3**). After correcting for multiple comparisons, we did not identify any HLA class I alleles that were significantly associated with spike-specific CD8+ T cell response frequencies after *two* vaccine doses, but the top allele after two vaccine doses was also A*02:03 (p=0.02; q=0.34; not shown).

To further explore the interaction with A*02:03, we used NetMHCpan-4.1 ^25^ to identify all 8-, 9-, and 10-amino acid spike epitopes that are predicted to bind to this allele or to the common and closely related A*02:01 allele, which showed no beneficial impact on CD8+ T cell responses after three vaccine doses (p=0.46, q=0.56; **Table S3**). In total, 29 strong binders (% Rank <0.5) and 56 weak binders (% Rank 0.5-2) were observed for A*02:03, compared to 20 strong binders and 58 weak binders for A*02:01 (**Table S4**). A cumulative view of strong binders, plotted by % Rank, is shown in **Figure 3A**. An analysis of 30 shared epitopes that displayed strong binding affinity for either allele further revealed that these epitopes were frequently predicted to bind more strongly to A*02:03 than A*02:01 (Wilcoxon matched pairs test, p=0.009) (**Figure 3B** and **Table S5**). Notably, seven weak-binding A*02:01 epitopes (all with % Ranks of ∼1 to 2) were predicted to bind A*02:03 strongly, while only one moderate-binding A*02:03 epitope (GLTVLPPLL, % Rank 0.69) was predicted to bind strongly to A*02:01 (% Rank 0.26) (**Figure 3B**). These results suggest that higher spike peptide affinity for A*02:03 contributes to enhanced CD8+ T cell responses following vaccination.

**Figure 3.**
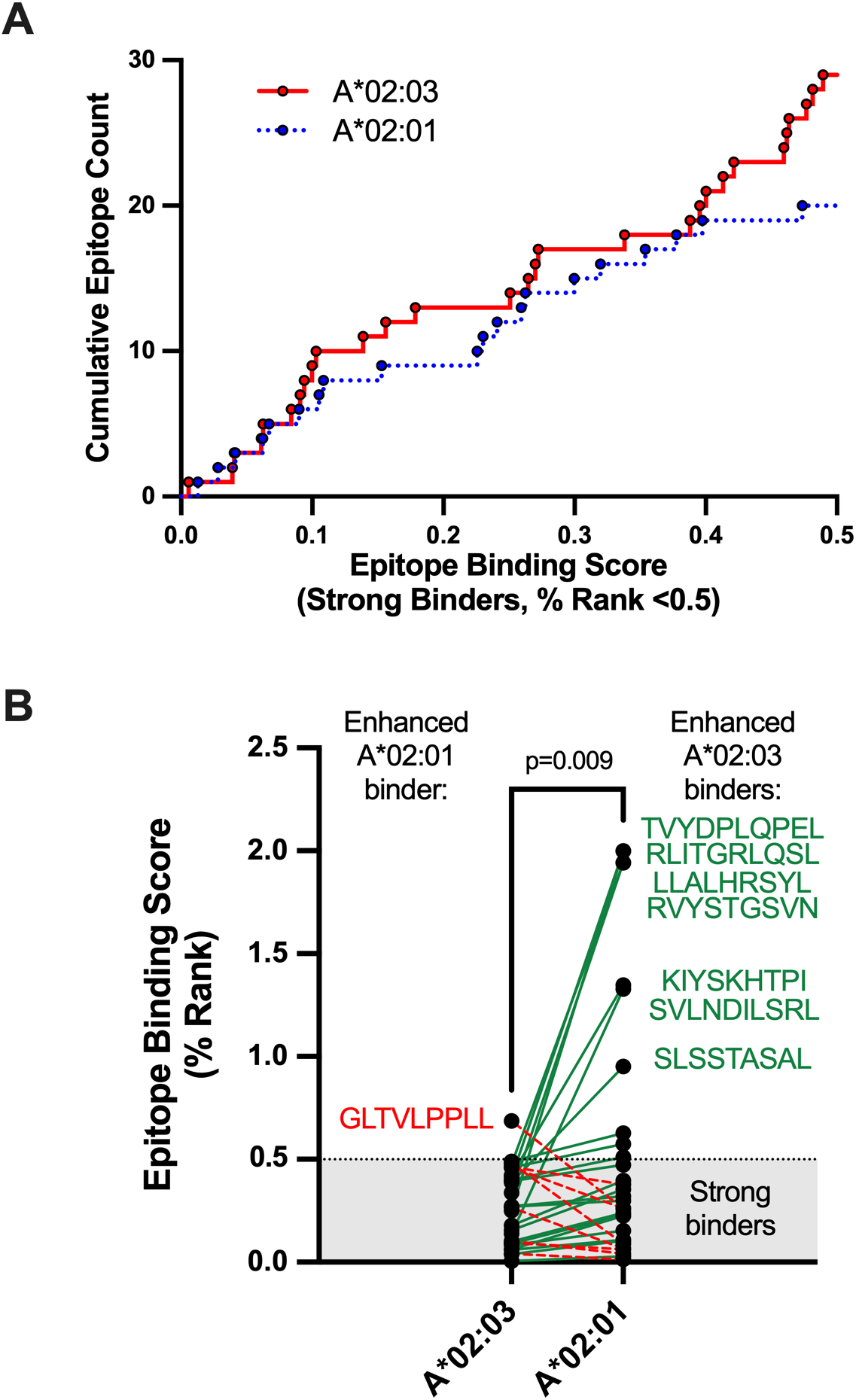
Comparison of HLA-A*02:01 and A*02:03 predicted strong-binding epitopes in SARS-CoV-2 spike. *Panel A:* Cumulative frequencies of predicted strong-binding epitopes in SARS-CoV-2 spike restricted by HLA-A*02:01 (blue line) and A*02:03. Epitope predictions were performed using NetMHCpan4.1 ^25^, which defines strong binders as those with % Ranks of 0.5 or lower. The lower the % Rank, the stronger the predicted binding. *Panel B:* Epitope binding ranks for 30 spike epitopes that were predicted to bind strongly to either A*02:01 or A*02:03. Epitopes that are predicted to bind more strongly to A*02:03 are linked by green lines, with seven epitopes that were predicted to have the most substantially enhanced A*02:03 binding listed in green next to their corresponding points. Epitopes that are predicted to bind more strongly to A*02:01 are linked by red dotted lines, with one epitope predicted to have the most substantially enhanced A*02:01 binding listed in red next to its corresponding point. The shaded area denotes the threshold for strong binders. P-value computed using the Wilcoxon matched pairs test.

### Breakthrough infection boosts CD4+ and CD8+ T-cell responses

Post-vaccination breakthrough SARS-CoV-2 infection, which was observed in 8 (20%) OA and 16 (32%) HCW, further boosted the percentage of both spike-specific CD4+ and CD8+ T-cells (p≤0.008 for all paired longitudinal comparisons; **Figures 4A, 4B**). The median percentage of spike-specific CD4+ T-cells increased to 0.73% (IQR 0.58-1.10) in OA and 0.86% (IQR 0.51-1.18) in HCW (between-group comparison p=0.78; **Figure 4A**) while the median percentage of spike-specific CD8+ T-cells increased to 0.98% (IQR 0.81-1.77) in OA and 1.13% (IQR 0.71-1.87) in HCW (between-group comparison p=0.66; **Figure 4B**). A full longitudinal representation of CD4+ and CD8+ T cell responses induced by vaccination and breakthrough infection, in all studied participants, is shown in **Figure S3.**

**Figure 4.**
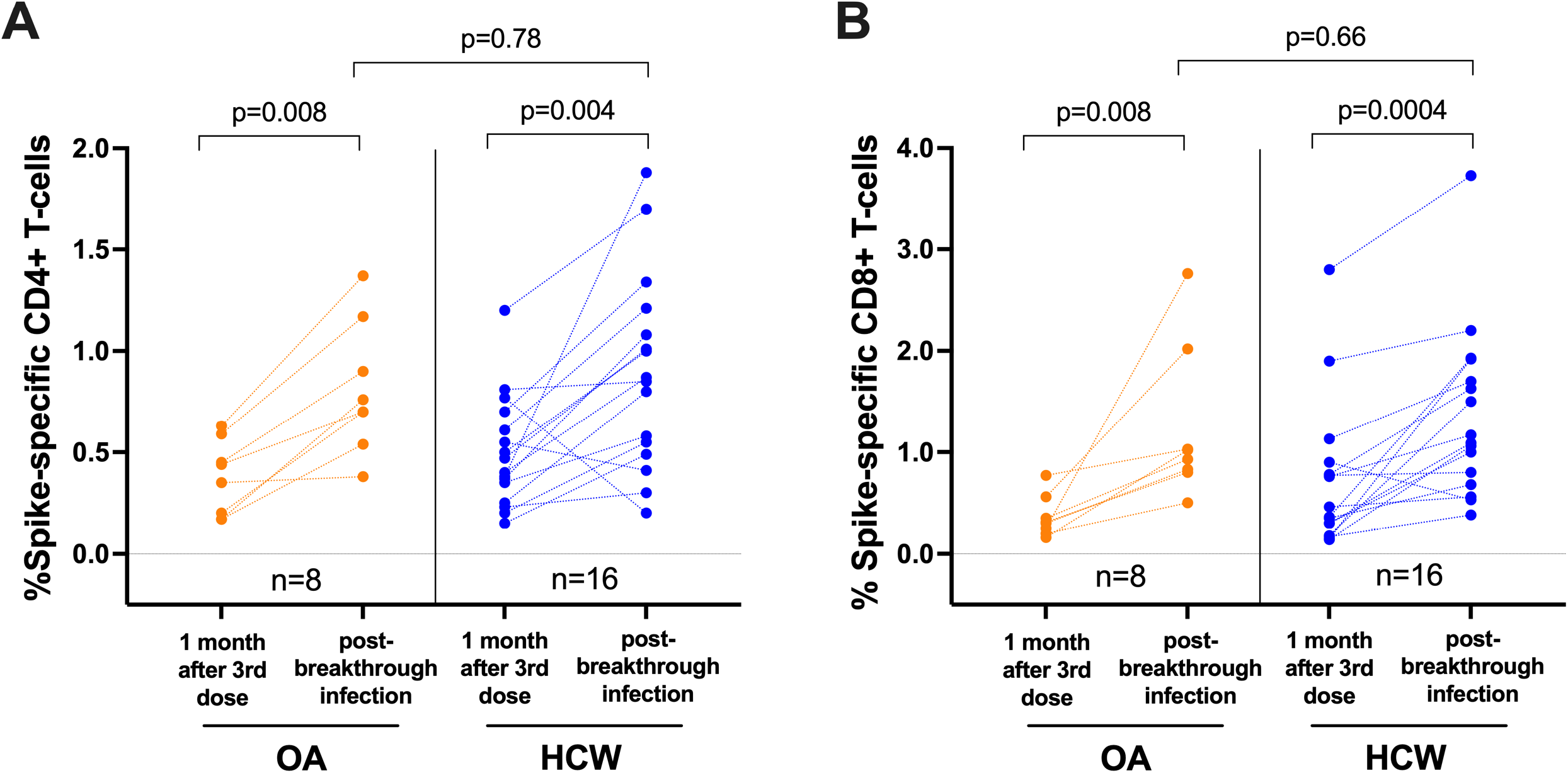
SARS-CoV-2 spike-specific T-cell frequencies in a subset of participants who experienced their first SARS-CoV-2 infection after receiving three vaccine doses. *Panel A*: Spike-specific CD4+ T-cell frequencies in a subset of 8 OA (orange) and 16 HCW (blue) after three-dose COVID-19 vaccination while participants were still COVID-19-naive, and after a subsequent breakthrough SARS-CoV-2 infection. The Mann-Whitney U-test was used for between-group comparisons and the Wilcoxon matched pairs test was used for longitudinal paired comparisons. P-values are not corrected for multiple comparisons. *Panel B:* Same as panel A, but for spike-specific CD8+ T-cell responses.

## DISCUSSION

We observed that the frequencies of SARS-CoV-2 spike-specific CD4+ and CD8+ T cells were not significantly different between older and younger adults after two doses of COVID-19 mRNA vaccine, and that these responses were enhanced similarly by a third vaccine dose regardless of age. Furthermore, both older and younger adults mounted robust T cell responses following a post-vaccine breakthrough infection. These results are consistent with a recent report showing that spike-specific T cell responses measured using an AIM assay were equivalent in older and younger adults after two doses of mRNA vaccine^21^, though another study reported impaired T cell effector function, including lower cytokine production, in older adults following two vaccine doses^13^. Prior studies have also found weaker T cell responses in older adults after only one vaccine dose^11, 21, 28^, but we did not examine this time point here. We also observed no association between CD4+ T cell responses and RBD-specific IgG antibodies after either two or three vaccine doses, suggesting that sufficient T cell help was provided to stimulate spike-specific B cell responses despite a wide range of antigen-specific CD4+ T cell frequencies. While our results do not completely rule out the possibility of age-related dysfunction in vaccine-induced T cells among older adults, our data suggest that the impact of any age-associated T cell impairment is likely to be modest after the second and third vaccine dose.

Our data also indicate that HLA class I alleles modulate the frequency of spike-specific CD8+ T cell responses following vaccination. Notably, participants expressing HLA-A*02:03 exhibited higher CD8+ T cell responses, while individuals expressing B*39:01, B*44:03, or C*07:01 exhibited lower responses (though these associations only reached statistical significance after three vaccine doses). Bioinformatics analyses predicted a higher number of spike peptides that bound with high affinity to HLA-A*02:03 compared to the closely related A*02:01 allele (which was not associated with a better vaccine response), suggesting that A*02:03 may elicit immunodominant responses to a broader array of vaccine-derived antigens. Several studies have reported associations between HLA genotype and COVID-19 severity or predicted COVID-19 vaccine immunogenicity^29–33^, which may underpin ethnicity-related differences in T cell responses that have been observed in some contexts^34, 35^. Indeed, A*02:03 occurs most frequently in individuals of South and East Asian ancestry, and therefore may contribute to enhanced T cell responses to COVID-19 vaccines in this population ^35^. Additional studies are needed to examine these HLA associations in greater detail.

This study has some limitations. While the AIM assay provides a very sensitive method to quantify spike-specific CD4+ and CD8+ T cells, we did not analyze T cell functions such as cytokine production or proliferation that could reveal differences in the effector activity of vaccine- or infection-induced responses. Since T cell responses were measured using peptide pools, we cannot comment on the relative dominance of individual spike epitopes (or host HLAs), nor potential changes in the distribution of these responses over time. Furthermore, we examined T cell responses using only peptides spanning the ancestral spike protein, which match the vaccine antigen but may not fully account for responses elicited against SARS-CoV-2 variants following breakthrough infection. Finally, we did not assess T cell responses after the first dose of COVID-19 mRNA vaccine, when age-related differences in immune cell frequency or function may be more apparent.

In summary, our study provides evidence that repeated exposure to SARS-CoV-2 spike antigen, either via vaccination or infection, induces steadily higher T cell frequencies in adults of all ages, at least up to 4 exposures. Importantly, our results also indicate that COVID-19-naive older adults mount robust cellular immune responses after two and three doses of COVID-19 mRNA vaccine, as well as after subsequent breakthrough infection, that are comparable in magnitude to those of younger adults. These results further underscore the benefits of COVID-19 vaccination in this population.

## Data Availability

All data produced in the present study are available upon reasonable request to the authors.

## AUTHOR CONTRIBUTIONS

ZLB and MAB led the study. HRL and YS coordinated the study. SD, RK, FM, SS, YS, RW, FY and EB processed specimens and collected data under the supervision of ZLB and MAB. SD, RL, MAB and ZLB analyzed data. MLD, DTH, JS, JSGM and MGR contributed to cohort establishment and/or SARS-CoV-2 serum antibody quantification. SD, ZLB and MAB wrote the manuscript. All authors contributed to manuscript review and editing.

## ACKNOWLEDGEMENTS

We thank the leadership and staff of Providence Health Care, St. Paul’s Hospital, the BC Centre for Excellence in HIV/AIDS, the Hope to Health Research and Innovation Centre, and Simon Fraser University for supporting this project. We also thank the participants, without whom this study would not have been possible.

## REFERENCES

1. Onder G, Rezza G, Brusaferro S. Case-Fatality Rate and Characteristics of Patients Dying in Relation to COVID-19 in Italy. JAMA 2020; 323(18): 1775–6.

2. Richardson S, Hirsch JS, Narasimhan M, et al. Presenting Characteristics, Comorbidities, and Outcomes Among 5700 Patients Hospitalized With COVID-19 in the New York City Area. JAMA 2020; 323(20): 2052–9.

3. Williamson EJ, Walker AJ, Bhaskaran K, et al. Factors associated with COVID-19-related death using OpenSAFELY. Nature 2020; 584(7821): 430–6.

4. Polack FP, Thomas SJ, Kitchin N, et al. Safety and Efficacy of the BNT162b2 mRNA Covid-19 Vaccine. N Engl J Med 2020; 383(27): 2603–15.

5. Goronzy JJ, Li G, Yang Z, Weyand CM. The janus head of T cell aging - autoimmunity and immunodeficiency. Front Immunol 2013; 4: 131.

6. Jefferson T, Rivetti D, Rivetti A, Rudin M, Di Pietrantonj C, Demicheli V. Efficacy and effectiveness of influenza vaccines in elderly people: a systematic review. Lancet 2005; 366(9492): 1165–74.

7. Nichol KL, Nordin JD, Nelson DB, Mullooly JP, Hak E. Effectiveness of influenza vaccine in the community-dwelling elderly. N Engl J Med 2007; 357(14): 1373–81.

8. Goronzy JJ, Weyand CM. Understanding immunosenescence to improve responses to vaccines. Nat Immunol 2013; 14(5): 428–36.

9. Goronzy JJ, Weyand CM. Mechanisms underlying T cell ageing. Nat Rev Immunol 2019; 19(9): 573–83.

10. Andrews N, Tessier E, Stowe J, et al. Duration of Protection against Mild and Severe Disease by Covid-19 Vaccines. N Engl J Med 2022; 386(4): 340–50.

11. Collier DA, Ferreira I, Kotagiri P, et al. Age-related immune response heterogeneity to SARS-CoV-2 vaccine BNT162b2. Nature 2021; 596(7872): 417–22.

12. Levin EG, Lustig Y, Cohen C, et al. Waning Immune Humoral Response to BNT162b2 Covid-19 Vaccine over 6 Months. N Engl J Med 2021; 385(24): e84.

13. Palacios-Pedrero MA, Jansen JM, Blume C, et al. Signs of immunosenescence correlate with poor outcome of mRNA COVID-19 vaccination in older adults. Nat Aging 2022; 2(10): 896–905.

14. Mwimanzi F, Lapointe HR, Cheung PK, et al. Impact of age and SARS-CoV-2 breakthrough infection on humoral immune responses after three doses of COVID-19 mRNA vaccine. Open Forum Infectious Diseases 2023; Accepted, In Press.

15. Mwimanzi F, Lapointe HR, Cheung PK, et al. Older Adults Mount Less Durable Humoral Responses to Two Doses of COVID-19 mRNA Vaccine but Strong Initial Responses to a Third Dose. J Infect Dis 2022; 226(6): 983–94.

16. US Food and Drug Administration. FDA Authorizes Booster Dose of Pfizer-BioNTech COVID-19 Vaccine for Certain Populations. fda.gov; 2021.

17. Bowman E. A CDC Panel Backs Booster Shots For Older Adults, A Step Toward Making Them Available. ‘National Public Radio, USA. 2021 23 Sept 2021.

18. Moss P. The T cell immune response against SARS-CoV-2. Nat Immunol 2022; 23(2): 186–93.

19. Painter MM, Mathew D, Goel RR, et al. Rapid induction of antigen-specific CD4(+) T cells is associated with coordinated humoral and cellular immunity to SARS-CoV-2 mRNA vaccination. Immunity 2021; 54(9): 2133–42 e3.

20. Sahin U, Muik A, Derhovanessian E, et al. COVID-19 vaccine BNT162b1 elicits human antibody and T(H)1 T cell responses. Nature 2020; 586(7830): 594–9.

21. Jo N, Hidaka Y, Kikuchi O, et al. Impaired CD4(+) T cell response in older adults is associated with reduced immunogenicity and reactogenicity of mRNA COVID-19 vaccination. Nat Aging 2023; 3(1): 82–92.

22. Brockman MA, Mwimanzi F, Lapointe HR, et al. Reduced Magnitude and Durability of Humoral Immune Responses to COVID-19 mRNA Vaccines Among Older Adults. J Infect Dis 2022; 225(7): 1129–40.

23. Brumme ZL, Brumme CJ, Chui C, et al. Effects of human leukocyte antigen class I genetic parameters on clinical outcomes and survival after initiation of highly active antiretroviral therapy. J Infect Dis 2007; 195(11): 1694–704.

24. Woods CK, Brumme CJ, Liu TF, et al. Automating HIV drug resistance genotyping with RECall, a freely accessible sequence analysis tool. J Clin Microbiol 2012; 50(6): 1936–42.

25. Reynisson B, Alvarez B, Paul S, Peters B, Nielsen M. NetMHCpan-4.1 and NetMHCIIpan-4.0: improved predictions of MHC antigen presentation by concurrent motif deconvolution and integration of MS MHC eluted ligand data. Nucleic Acids Res 2020; 48(W1): W449–W54.

26. Storey JD, Tibshirani R. Statistical significance for genomewide studies. Proc Natl Acad Sci U S A 2003; 100(16): 9440–5.

27. BC Centre for Disease Control. Weekly update on Variants of Concern. 2022. http://www.bccdc.ca/health-info/diseases-conditions/covid-19/data.

28. Muller L, Andree M, Moskorz W, et al. Age-dependent Immune Response to the Biontech/Pfizer BNT162b2 Coronavirus Disease 2019 Vaccination. Clin Infect Dis 2021; 73(11): 2065–72.

29. Bose T, Pant N, Pinna NK, Bhar S, Dutta A, Mande SS. Does immune recognition of SARS-CoV2 epitopes vary between different ethnic groups? Virus Res 2021; 305: 198579.

30. Rao V, Chandra N. In-silico study of influence of HLA heterogeneity on CTL responses across ethnicities to SARS-CoV-2. Hum Immunol 2022; 83(12): 797–802.

31. Bertinetto FE, Magistroni P, Mazzola GA, et al. The humoral and cellular response to mRNA SARS-CoV-2 vaccine is influenced by HLA polymorphisms. HLA 2023.

32. Srivastava A, Hollenbach JA. The immunogenetics of COVID-19. Immunogenetics 2023; 75(3): 309–20.

33. Sacco K, Castagnoli R, Vakkilainen S, et al. Immunopathological signatures in multisystem inflammatory syndrome in children and pediatric COVID-19. Nat Med 2022; 28(5): 1050–62.

34. Smith M, Abdesselem HB, Mullins M, et al. Age, Disease Severity and Ethnicity Influence Humoral Responses in a Multi-Ethnic COVID-19 Cohort. Viruses 2021; 13(5).

35. Martin CA, Nazareth J, Jarkhi A, et al. Ethnic differences in cellular and humoral immune responses to SARS-CoV-2 vaccination in UK healthcare workers: a cross-sectional analysis. EClinicalMedicine 2023; 58: 101926.

