## Supplemental Figures for "Dynamics of T-cell responses following COVID-19 mRNA vaccination and breakthrough infection in older adults"

**Figure S1**

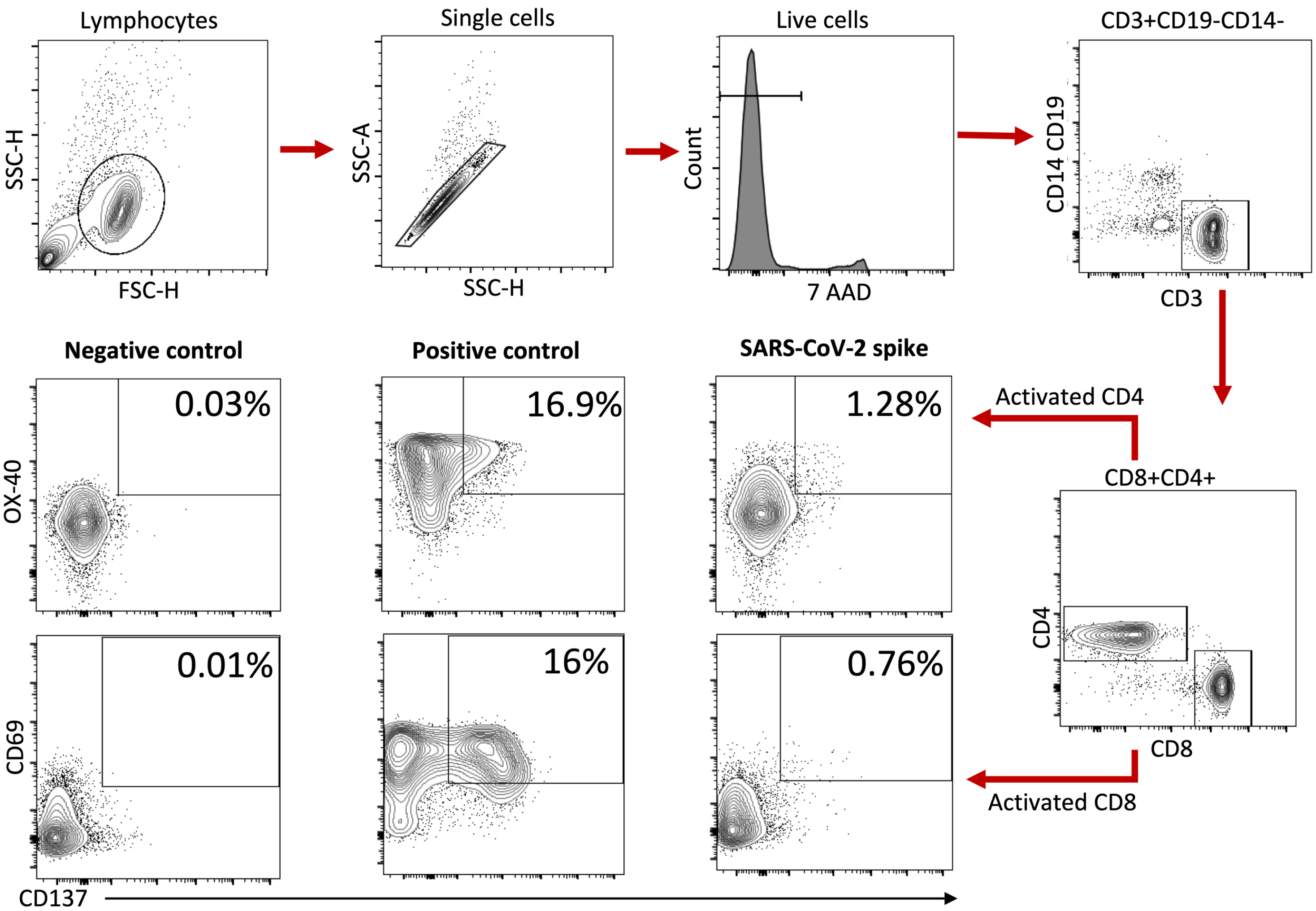


**Figure S1. Gating strategy for quantification of SARS-CoV-2 Spike-specific CD4+ and CD8+ T-cells using an activation induced marker (AIM) assay.**

Figure S2

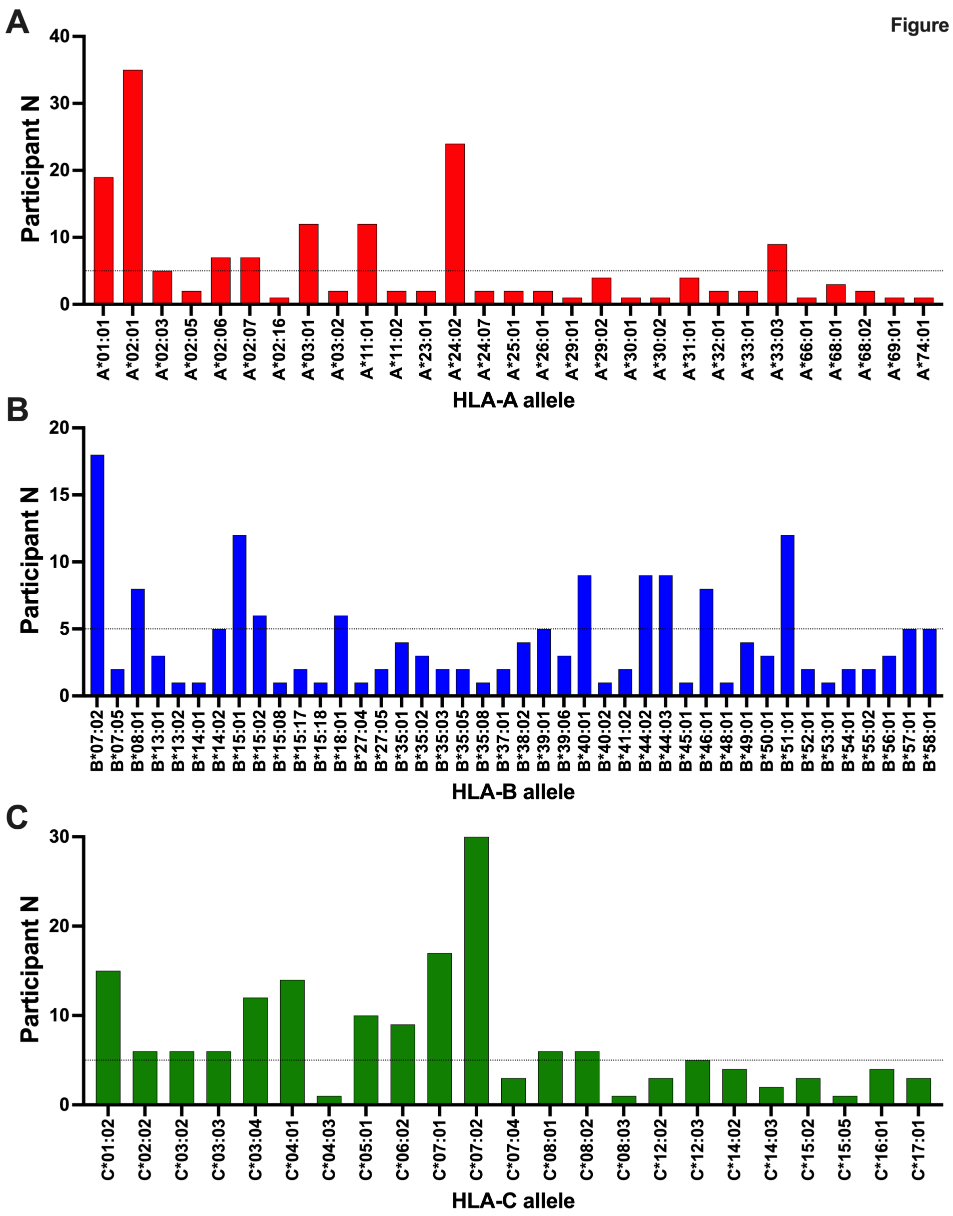


**Figure S2. HLA-A, HLA-B and HLA-C allele frequencies, reported as the number of participants expressing each allele in the overall cohort.** Alleles observed a minimum of 5 times, shown by the dotted line, were assessed for their relationship with SARS-CoV-2 Spike-specific CD8+ T cell frequencies post-vaccination.

Figure S3

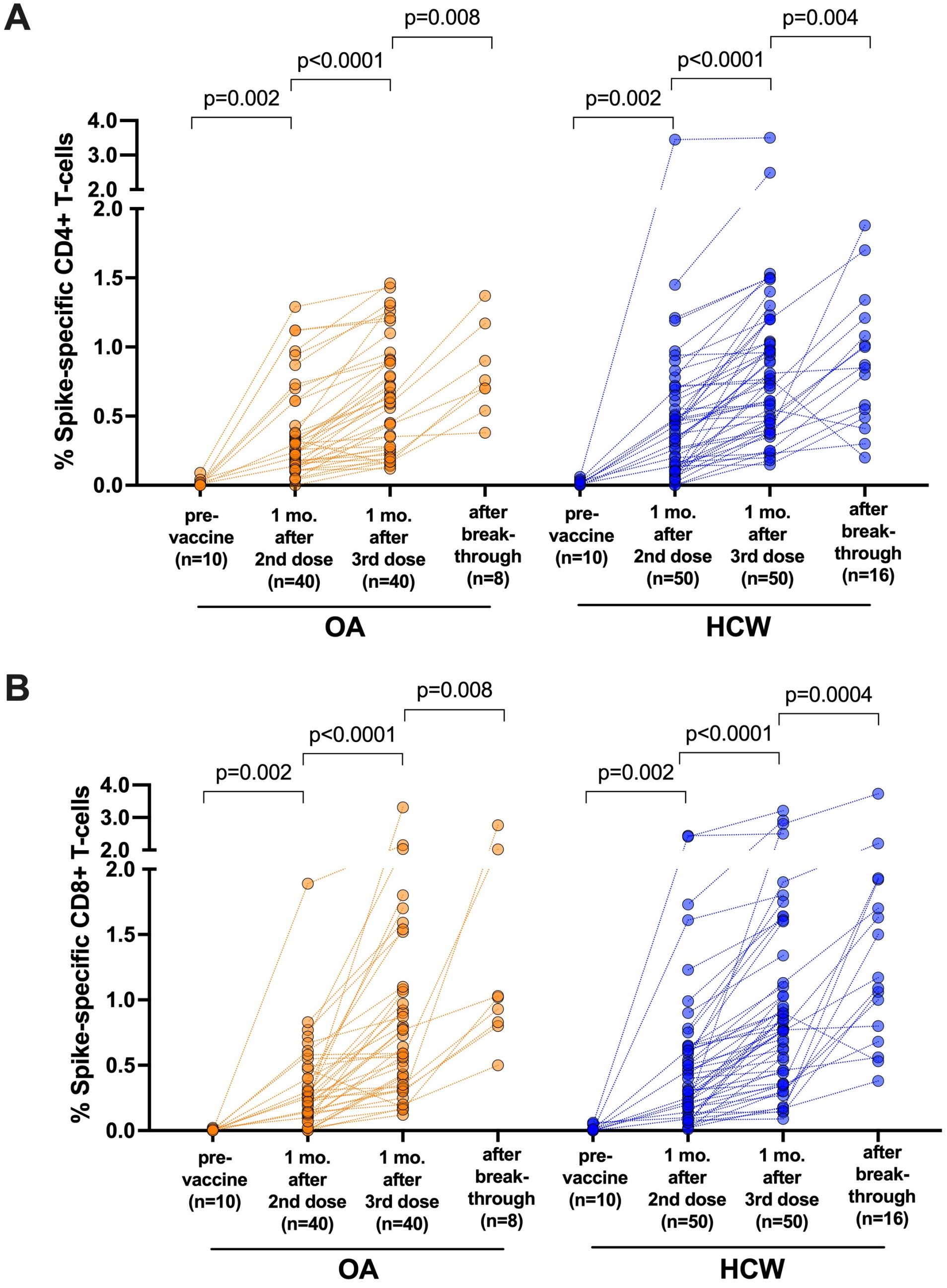


**Figure S3. Longitudinal view of SARS-CoV-2 Spike-specific T-cell frequencies before and after vaccination and subsequent breakthrough infection.** These are the same data as shown in Figure 1, but plotted longitudinally by participant. *Panel A:* CD4+ T-cell frequencies. *Panel B:* CD8+ T-cell frequencies. Older Adults (OA) are in orange; younger Health Care Workers (HCW) are in blue. P-values are calculated using the Wilcoxon matched pairs test.
