## Supplemental Tables for "Dynamics of T-cell responses following COVID-19 mRNA vaccination and breakthrough infection in older adults"

**Table S1: Multivariable zero-inflated beta regressions exploring the relationship between sociodemographic, health and vaccine-related variables on Spike-specific T-cell responses following two COVID-19 mRNA vaccine doses**

| Variable | Outcome measure | | | | | |
|---|---|---|---|---|---|---|
| | % Spike-responsive CD4+ T-cells after two vaccine doses | | | % Spike-responsive CD8+ T-cells after two vaccine doses | | |
| | **Estimate** | **Standard error** | **p-value** | **Estimate** | **Standard error** | **p-value** |
| Age (per year) | 0.0038 | 0.0042 | 0.36 | -0.00035 | 0.0047 | 0.94 |
| Male sex | 0.15 | 0.16 | 0.37 | 0.057 | 0.18 | 0.75 |
| White ethnicity | -0.38 | 0.17 | **0.03** | -0.0021 | 0.18 | 0.99 |
| # Health conditions (per additional)[a] | 0.0014 | 0.077 | 0.99 | 0.033 | 0.089 | 0.71 |
| mRNA-1273-containing initial vaccine regimen[b] | -0.041 | 0.25 | 0.87 | -0.013 | 0.26 | 0.96 |
| Days between 1st and 2nd vaccine dose | 0.0038 | 0.0034 | 0.27 | 0.0040 | 0.0037 | 0.28 |

[a]Defined as: hypertension, diabetes, asthma, obesity (defined as having a body mass index ≥30), chronic diseases of lung, liver, kidney, heart or blood, cancer, and immunosuppression due to chronic conditions or medication.
[b]Defined as two doses of mRNA-1273 or a heterologous regimen consisting of one dose each of mRNA-1273 and BNT162b2

**Table S2: Multivariable zero-inflated beta regressions exploring the relationship between sociodemographic, health and vaccine-related variables on Spike-specific T-cell responses following three COVID-19 mRNA vaccine doses**

| Variable | Outcome measure | | | | | |
|---|---|---|---|---|---|---|
| | **% Spike-responsive CD4+ T-cells after three vaccine doses** | | | **% Spike-responsive CD8+ T-cells after three vaccine doses** | | |
| | **Estimate** | **Standard error** | **p-value** | **Estimate** | **Standard error** | **p-value** |
| Age (per year) | -0.004 | 0.0037 | 0.26 | 0.0015 | 0.0043 | 0.72 |
| Male sex | 0.23 | 0.10 | **0.03** | 0.070 | 0.13 | 0.58 |
| White ethnicity | -0.18 | 0.10 | 0.08 | -0.11 | 0.12 | 0.36 |
| # Health conditions (per additional)[a] | 0.13 | 0.046 | **0.006** | -0.0076 | 0.057 | 0.89 |
| mRNA-1273-containing initial vaccine regimen[b] | 0.031 | 0.15 | 0.84 | 0.074 | 0.19 | 0.70 |
| Days between 1st and 2nd vaccine dose | 0.0037 | 0.0032 | 0.25 | 0.0029 | 0.0039 | 0.46 |
| mRNA-1273 as third vaccine dose[c] | 0.23 | 0.11 | **0.03** | 0.039 | 0.12 | 0.74 |
| Days between 2nd and 3rd vaccine dose | 0.0016 | 0.0021 | 0.43 | -0.00018 | 0.0024 | 0.94 |
| % Spike responsive T-cells after two doses (per 1% increment)[d] | 0.69 | 0.062 | **$2x10^{-16}$** | 0.77 | 0.086 | **$1x10^{-13}$** |

[a]Defined as: hypertension, diabetes, asthma, obesity (defined as having a body mass index ≥30), chronic diseases of lung, liver, kidney, heart or blood, cancer, and immunosuppression due to chronic conditions or medication.
[b]Defined as two doses of mRNA-1273, or a heterologous regimen consisting of one dose each of mRNA-1273 and BNT162b2 (vs. a reference group defined as two doses of BNT162b2).
[c]Reference group defined as a BNT162b2 third vaccine dose
[d]Post-second dose % CD4+ and CD8+ T-cell responsiveness values were used for the post-third dose CD4+ and CD8+ T-cell models, respectively.

**Table S3. Associations between HLA class I alleles and SARS-CoV-2 spike-specific CD8+ T-cell responses after three COVID-19 mRNA vaccine doses**

| Allele | N with[a] | N without | median with[b] | median without | p-value[c] | q-value |
|---|---|---|---|---|---|---|
| **B*39:01** | **5** | **85** | **0.27** | **0.77** | **0.004** | **0.08** |
| **A*02:03** | **5** | **85** | **2.50** | **0.73** | **0.01** | **0.08** |
| **B*44:03** | **9** | **81** | **0.46** | **0.78** | **0.01** | **0.08** |
| **C*07:01** | **17** | **73** | **0.57** | **0.78** | **0.02** | **0.13** |
| C*12:03 | 5 | 85 | 1.70 | 0.76 | 0.06 | 0.27 |
| B*57:01 | 5 | 85 | 1.10 | 0.76 | 0.11 | 0.40 |
| B*46:01 | 8 | 82 | 0.93 | 0.75 | 0.14 | 0.41 |
| C*01:02 | 15 | 75 | 0.87 | 0.73 | 0.18 | 0.41 |
| B*51:01 | 12 | 78 | 1.21 | 0.76 | 0.18 | 0.41 |
| C*02:02 | 6 | 84 | 0.49 | 0.77 | 0.20 | 0.41 |
| B*14:02 | 5 | 85 | 0.78 | 0.73 | 0.20 | 0.41 |
| B*08:01 | 8 | 82 | 0.60 | 0.77 | 0.21 | 0.41 |
| A*03:01 | 12 | 78 | 0.64 | 0.77 | 0.29 | 0.49 |
| B*58:01 | 5 | 85 | 0.90 | 0.76 | 0.31 | 0.49 |
| A*24:02 | 24 | 66 | 0.74 | 0.77 | 0.32 | 0.49 |
| C*03:02 | 6 | 84 | 0.88 | 0.75 | 0.36 | 0.51 |
| A*11:01 | 12 | 78 | 0.50 | 0.77 | 0.39 | 0.52 |
| A*02:01 | 35 | 55 | 0.77 | 0.73 | 0.46 | 0.56 |
| A*02:07 | 7 | 83 | 0.90 | 0.76 | 0.48 | 0.56 |
| A*01:01 | 19 | 71 | 0.62 | 0.77 | 0.50 | 0.56 |
| C*07:02 | 30 | 60 | 0.77 | 0.77 | 0.52 | 0.56 |
| C*03:04 | 12 | 78 | 0.85 | 0.75 | 0.57 | 0.56 |
| C*08:02 | 6 | 84 | 0.78 | 0.75 | 0.58 | 0.56 |
| B*15:01 | 12 | 78 | 0.77 | 0.76 | 0.59 | 0.56 |
| B*18:01 | 6 | 84 | 0.73 | 0.77 | 0.67 | 0.56 |
| B*07:02 | 18 | 72 | 0.77 | 0.77 | 0.70 | 0.56 |
| B*15:02 | 6 | 84 | 0.79 | 0.77 | 0.72 | 0.56 |
| C*08:01 | 6 | 84 | 0.79 | 0.77 | 0.72 | 0.56 |
| B*40:01 | 9 | 81 | 0.90 | 0.76 | 0.72 | 0.56 |

| | | | | | | |
|---|---|---|---|---|---|---|
| A*33:03 | 9 | 81 | 0.76 | 0.77 | 0.78 | 0.57 |
| B*44:02 | 9 | 81 | 0.85 | 0.76 | 0.80 | 0.57 |
| C*05:01 | 10 | 80 | 0.75 | 0.77 | 0.82 | 0.57 |
| C*04:01 | 14 | 76 | 0.77 | 0.77 | 0.82 | 0.57 |
| C*06:02 | 9 | 81 | 0.59 | 0.77 | 0.87 | 0.57 |
| C*03:03 | 6 | 84 | 0.77 | 0.76 | 0.91 | 0.57 |
| A*02:06 | 7 | 83 | 0.87 | 0.76 | 0.91 | 0.57 |

Results are sorted by statistical significance, with bold text denoting associations that meet the significance threshold after correction for multiple comparisons.
[a] N with/without: the N of participants expressing or not expressing the allele, respectively. Analysis was restricted to alleles expressed by a minimum of 5 participants.
[b] median with/without: the median spike-specific CD8+ T-cell frequencies after the third vaccine dose, in participants expressing or not expressing the allele, respectively
[c] p-values are calculated using the Mann-Whitney U test.

**Table S4. HLA A*02:01 and A*02:03 binding epitopes in SARS-CoV-2 spike (predicted using NetMHCpan v4.1)**

| HLA | core | icore | EL_score | EL_rank | Cumulative rank | Strong Binder (SB) |
|---|---|---|---|---|---|---|
| HLA-A02:01 | YLQPRTFLL | YLQPRTFLL | 0.9712 | 0.0129 | 1 | SB |
| HLA-A02:01 | VLNDILSRL | VLNDILSRL | 0.9385 | 0.028 | 2 | SB |
| HLA-A02:01 | TLDSKTQSL | TLDSKTQSL | 0.915 | 0.0414 | 3 | SB |
| HLA-A02:01 | RLQSLQTYV | RLQSLQTYV | 0.8738 | 0.0622 | 4 | SB |
| HLA-A02:01 | KIADYNYKL | KIADYNYKL | 0.8646 | 0.0671 | 5 | SB |
| HLA-A02:01 | RLDKVEAEV | RLDKVEAEV | 0.825 | 0.0899 | 6 | SB |
| HLA-A02:01 | LLFNKVTLA | LLFNKVTLA | 0.8035 | 0.1053 | 7 | SB |
| HLA-A02:01 | HLMSFPQSA | HLMSFPQSA | 0.7985 | 0.1085 | 8 | SB |
| HLA-A02:01 | VVFLHVTYV | VVFLHVTYV | 0.7417 | 0.1528 | 9 | SB |
| HLA-A02:01 | ALNTLVKQL | ALNTLVKQL | 0.6574 | 0.2258 | 10 | SB |
| HLA-A02:01 | RLNEVAKNL | RLNEVAKNL | 0.6527 | 0.2303 | 11 | SB |
| HLA-A02:01 | FIAGLIAIV | FIAGLIAIV | 0.6414 | 0.2409 | 12 | SB |
| HLA-A02:01 | GLTVLPPLL | GLTVLPPLL | 0.6222 | 0.2593 | 13 | SB |
| HLA-A02:01 | NLNESLIDL | NLNESLIDL | 0.6189 | 0.2624 | 14 | SB |
| HLA-A02:01 | SIIAYTMSL | SIIAYTMSL | 0.58 | 0.2998 | 15 | SB |
| HLA-A02:01 | KLPDDFTGV | KLPDDFTGCV | 0.5628 | 0.3198 | 16 | SB |
| HLA-A02:01 | KLNDLFTNV | KLNDLCFTNV | 0.5334 | 0.3539 | 17 | SB |
| HLA-A02:01 | FLLHAPATV | FELLHAPATV | 0.5132 | 0.3775 | 18 | SB |
| HLA-A02:01 | VLYENQKLI | VLYENQKLI | 0.4959 | 0.3975 | 19 | SB |
| HLA-A02:01 | YLMSFPQSA | YHLMSFPQSA | 0.4392 | 0.4737 | 20 | SB |
| HLA-A02:01 | SLNDILSRL | SVLNDILSRL | 0.4151 | 0.5079 | 21 | |
| HLA-A02:01 | KLQDVVNQA | KLQDVVNQNA | 0.4133 | 0.511 | 22 | |
| HLA-A02:01 | RLITGLQSL | RLITGRLQSL | 0.41 | 0.5166 | 23 | |
| HLA-A02:01 | TVYDPLPEL | TVYDPLQPEL | 0.4092 | 0.5181 | 24 | |
| HLA-A02:01 | SLID-LQEL | SLIDLQEL | 0.3954 | 0.5417 | 25 | |
| HLA-A02:01 | FTISVTTEI | FTISVTTEI | 0.3759 | 0.575 | 26 | |
| HLA-A02:01 | FQFCNDPFL | FQFCNDPFL | 0.3691 | 0.5866 | 27 | |

| | | | | | |
|---|---|---|---|---|---|
| HLA-A02:01 | VLSFELLHA | VLSFELLHA | 0.3507 | 0.627 | 28 |
| HLA-A02:01 | ELLHAPATV | ELLHAPATV | 0.3364 | 0.6636 | 29 |
| HLA-A02:01 | KLPDDFTGC | KLPDDFTGC | 0.3349 | 0.6675 | 30 |
| HLA-A02:01 | TLDSKTQSL | TLDSKTQSLL | 0.2705 | 0.8537 | 31 |
| HLA-A02:01 | TLDSKTQSL | TTLDSKTQSL | 0.252 | 0.9172 | 32 |
| HLA-A02:01 | SLSSTASAL | SLSSTASAL | 0.2434 | 0.9509 | 33 |
| HLA-A02:01 | KLNESLIDL | KNLNESLIDL | 0.2428 | 0.9529 | 34 |
| HLA-A02:01 | SLQTVTQQL | SLQTYVTQQL | 0.2318 | 0.9958 | 35 |
| HLA-A02:01 | NTQEVFAQV | NTQEVFAQV | 0.2274 | 1.0149 | 36 |
| HLA-A02:01 | VTWFHAIHV | VTWFHAIHV | 0.2214 | 1.0425 | 37 |
| HLA-A02:01 | SVTTEILPV | SVTTEILPV | 0.2167 | 1.0636 | 38 |
| HLA-A02:01 | FCNDPFLGV | FCNDPFLGV | 0.2133 | 1.0794 | 39 |
| HLA-A02:01 | FVSNGTWFV | FVSNGTHWFV | 0.212 | 1.0853 | 40 |
| HLA-A02:01 | GLQSLQTYV | GRLQSLQTYV | 0.2107 | 1.091 | 41 |
| HLA-A02:01 | VYDPLQPEL | VYDPLQPEL | 0.1966 | 1.1573 | 42 |
| HLA-A02:01 | YQPYRVVVL | YQPYRVVVL | 0.189 | 1.1933 | 43 |
| HLA-A02:01 | YLQPRTF-L | YLQPRTFL | 0.1884 | 1.1964 | 44 |
| HLA-A02:01 | FL-PFFSNV | FLPFFSNV | 0.1844 | 1.2166 | 45 |
| HLA-A02:01 | LLFNKV-TL | LLFNKVTL | 0.1765 | 1.257 | 46 |
| HLA-A02:01 | MIAQYTSAL | MIAQYTSAL | 0.1764 | 1.2577 | 47 |
| HLA-A02:01 | SLDKVEAEV | SRLDKVEAEV | 0.1753 | 1.2633 | 48 |
| HLA-A02:01 | ALGKLQDVV | ALGKLQDVV | 0.1733 | 1.2734 | 49 |
| HLA-A02:01 | VLYE-NQKL | VLYENQKL | 0.1725 | 1.2778 | 50 |
| HLA-A02:01 | KQIYKTPPI | KQIYKTPPI | 0.1717 | 1.2818 | 51 |
| HLA-A02:01 | RVYSTGSNV | RVYSTGSNV | 0.1643 | 1.3286 | 52 |
| HLA-A02:01 | KIYSKHTPI | KIYSKHTPI | 0.1618 | 1.3468 | 53 |
| HLA-A02:01 | VLYQGVNCT | VLYQGVNCT | 0.1603 | 1.3579 | 54 |
| HLA-A02:01 | LQIPFAMQM | LQIPFAMQM | 0.1573 | 1.3801 | 55 |
| HLA-A02:01 | ELDSFKEEL | ELDSFKEEL | 0.149 | 1.434 | 56 |
| HLA-A02:01 | ALIPFAMQM | ALQIPFAMQM | 0.146 | 1.4515 | 57 |

| | | | | | | |
|---|---|---|---|---|---|---|
| HLA-A02:01 | FLV-LLPLV | FLVLLPLV | 0.1408 | 1.4827 | 58 | |
| HLA-A02:01 | GLQPRTFLL | GYLQPRTFLL | 0.139 | 1.4934 | 59 | |
| HLA-A02:01 | GINASVVNI | GINASVVNI | 0.1372 | 1.5054 | 60 | |
| HLA-A02:01 | FIEDLLFKV | FIEDLLFNKV | 0.1371 | 1.5062 | 61 | |
| HLA-A02:01 | AQLTPTWRV | ADQLTPTWRV | 0.1315 | 1.5466 | 62 | |
| HLA-A02:01 | ILDITPCSF | ILDITPCSF | 0.1313 | 1.5481 | 63 | |
| HLA-A02:01 | YTNSFTRGV | YTNSFTRGV | 0.1216 | 1.6254 | 64 | |
| HLA-A02:01 | VLYENQLIA | VLYENQKLIA | 0.1203 | 1.6393 | 65 | |
| HLA-A02:01 | AIPNFTISV | AIPTNFTISV | 0.1195 | 1.6483 | 66 | |
| HLA-A02:01 | VLHSQDLFL | VLHSTQDLFL | 0.1166 | 1.6786 | 67 | |
| HLA-A02:01 | PLVDLPIGI | PLVDLPIGI | 0.1158 | 1.6868 | 68 | |
| HLA-A02:01 | EQDKNTQEV | EQDKNTQEV | 0.1128 | 1.721 | 69 | |
| HLA-A02:01 | ALLAGTITS | ALLAGTITS | 0.1126 | 1.7229 | 70 | |
| HLA-A02:01 | YILGFIAGL | YIWLGFIAGL | 0.1079 | 1.7765 | 71 | |
| HLA-A02:01 | FLHVTYVPA | FLHVTYVPA | 0.0988 | 1.9043 | 72 | |
| HLA-A02:01 | LITGRLQSL | LITGRLQSL | 0.0982 | 1.9126 | 73 | |
| HLA-A02:01 | LLALHRSYL | LLALHRSYL | 0.0959 | 1.942 | 74 | |
| HLA-A02:01 | GIADYNYKL | GKIADYNYKL | 0.0947 | 1.9582 | 75 | |
| HLA-A02:01 | QLNRALTGI | QLNRALTGI | 0.0936 | 1.9735 | 76 | |
| HLA-A02:01 | YVTQQLIRA | YVTQQLIRA | 0.0917 | 1.9986 | 77 | |
| HLA-A02:03 | VLNDILSRL | VLNDILSRL | 0.9732 | 0.0058 | 1 | SB |
| HLA-A02:03 | LLFNKVTLA | LLFNKVTLA | 0.8901 | 0.0391 | 2 | SB |
| HLA-A02:03 | YLQPRTFLL | YLQPRTFLL | 0.8867 | 0.0406 | 3 | SB |
| HLA-A02:03 | HLMSFPQSA | HLMSFPQSA | 0.8455 | 0.0611 | 4 | SB |
| HLA-A02:03 | ALNTLVKQL | ALNTLVKQL | 0.8428 | 0.0626 | 5 | SB |
| HLA-A02:03 | VVFLHVTYV | VVFLHVTYV | 0.8038 | 0.084 | 6 | SB |
| HLA-A02:03 | RLNEVAKNL | RLNEVAKNL | 0.7887 | 0.0907 | 7 | SB |
| HLA-A02:03 | RLQSLQTYV | RLQSLQTYV | 0.7798 | 0.0939 | 8 | SB |
| HLA-A02:03 | FIAGLIAIV | FIAGLIAIV | 0.7635 | 0.0997 | 9 | SB |
| HLA-A02:03 | TLDSKTQSL | TLDSKTQSL | 0.7588 | 0.103 | 10 | SB |

| | | | | | | |
|---|---|---|---|---|---|---|
| HLA-A02:03 | SLNDILSRL | SVLNDILSRL | 0.7044 | 0.1387 | 11 | SB |
| HLA-A02:03 | KLNDLFTNV | KLNDLCFTNV | 0.6752 | 0.156 | 12 | SB |
| HLA-A02:03 | VLYENQKLI | VLYENQKLI | 0.649 | 0.1785 | 13 | SB |
| HLA-A02:03 | RLIGRLQSL | RLITGRLQSL | 0.5762 | 0.2509 | 14 | SB |
| HLA-A02:03 | KIADYNYKL | KIADYNYKL | 0.5631 | 0.2647 | 15 | SB |
| HLA-A02:03 | SIIAYTMSL | SIIAYTMSL | 0.5582 | 0.27 | 16 | SB |
| HLA-A02:03 | KLPDDFTGV | KLPDDFTGCV | 0.5561 | 0.2722 | 17 | SB |
| HLA-A02:03 | RVYSTGSNV | RVYSTGSNV | 0.5005 | 0.3381 | 18 | SB |
| HLA-A02:03 | SLSSTASAL | SLSSTASAL | 0.4618 | 0.3881 | 19 | SB |
| HLA-A02:03 | YLMSFPQSA | YHLMSFPQSA | 0.4561 | 0.3955 | 20 | SB |
| HLA-A02:03 | KLQDVVQNA | KLQDVVNQNA | 0.4524 | 0.4003 | 21 | SB |
| HLA-A02:03 | TVYDPLQEL | TVYDPLQPEL | 0.4446 | 0.4133 | 22 | SB |
| HLA-A02:03 | KIYSKHTPI | KIYSKHTPI | 0.4398 | 0.4213 | 23 | SB |
| HLA-A02:03 | FTISVTTEI | FTISVTTEI | 0.4168 | 0.4596 | 24 | SB |
| HLA-A02:03 | FLLHAPATV | FELLHAPATV | 0.4156 | 0.4617 | 25 | SB |
| HLA-A02:03 | NLNESLIDL | NLNESLIDL | 0.4146 | 0.4634 | 26 | SB |
| HLA-A02:03 | LLALHRSYL | LLALHRSYL | 0.4066 | 0.4767 | 27 | SB |
| HLA-A02:03 | RLDKVEAEV | RLDKVEAEV | 0.4037 | 0.4814 | 28 | SB |
| HLA-A02:03 | VLSFELLHA | VLSFELLHA | 0.399 | 0.4893 | 29 | SB |
| HLA-A02:03 | QLNRALTGI | QLNRALTGI | 0.3819 | 0.5257 | 30 | |
| HLA-A02:03 | MIAQYTSAL | MIAQYTSAL | 0.3605 | 0.577 | 31 | |
| HLA-A02:03 | YTNSFTRGV | YTNSFTRGV | 0.346 | 0.6123 | 32 | |
| HLA-A02:03 | GINASVVNI | GINASVVNI | 0.3306 | 0.6516 | 33 | |
| HLA-A02:03 | ELLHAPATV | ELLHAPATV | 0.3173 | 0.6855 | 34 | |
| HLA-A02:03 | GLTVLPPLL | GLTVLPPLL | 0.3164 | 0.6876 | 35 | |
| HLA-A02:03 | SLID-LQEL | SLIDLQEL | 0.3022 | 0.7326 | 36 | |
| HLA-A02:03 | FL-PFFSNV | FLPFFSNV | 0.2951 | 0.7573 | 37 | |
| HLA-A02:03 | KLPDDFTGC | KLPDDFTGC | 0.2798 | 0.8117 | 38 | |
| HLA-A02:03 | LITGRLQSL | LITGRLQSL | 0.2672 | 0.8597 | 39 | |
| HLA-A02:03 | SVTTEILPV | SVTTEILPV | 0.2468 | 0.9406 | 40 | |

| | | | | | |
|---|---|---|---|---|---|
| HLA-A02:03 | SLQTVTQQL | SLQTYVTQQL | 0.2465 | 0.9416 | 41 |
| HLA-A02:03 | VLYQGVNCT | VLYQGVNCT | 0.2414 | 0.9626 | 42 |
| HLA-A02:03 | NTQEVFAQV | NTQEVFAQV | 0.2378 | 0.9773 | 43 |
| HLA-A02:03 | FVSNGTHFV | FVSNGTHWFV | 0.2339 | 0.9934 | 44 |
| HLA-A02:03 | KLINQFNSA | KLIANQFNSA | 0.2302 | 1.0095 | 45 |
| HLA-A02:03 | YQPYRVVVL | YQPYRVVVL | 0.2263 | 1.0279 | 46 |
| HLA-A02:03 | FLHVTYVPA | FLHVTYVPA | 0.2181 | 1.0662 | 47 |
| HLA-A02:03 | AQKFNGLTV | AQKFNGLTV | 0.2162 | 1.0747 | 48 |
| HLA-A02:03 | GVFLHVTYV | GVVFLHVTYV | 0.2084 | 1.1134 | 49 |
| HLA-A02:03 | KQIYKTPPI | KQIYKTPPI | 0.196 | 1.1821 | 50 |
| HLA-A02:03 | FCNDPFLGV | FCNDPFLGV | 0.1946 | 1.1896 | 51 |
| HLA-A02:03 | VLYENQKLA | VLYENQKLIA | 0.1931 | 1.1975 | 52 |
| HLA-A02:03 | GLQSLQTYV | GRLQSLQTYV | 0.1879 | 1.2252 | 53 |
| HLA-A02:03 | LLFNKVT-L | LLFNKVTL | 0.181 | 1.2617 | 54 |
| HLA-A02:03 | LIVNNATNV | LIVNNATNV | 0.1603 | 1.4034 | 55 |
| HLA-A02:03 | VVIGIVNTV | VVIGIVNNTV | 0.1552 | 1.4509 | 56 |
| HLA-A02:03 | VLYEN-QKL | VLYENQKL | 0.1541 | 1.4612 | 57 |
| HLA-A02:03 | AIPNFTISV | AIPTNFTISV | 0.1525 | 1.4752 | 58 |
| HLA-A02:03 | IVNNATNVV | IVNNATNVV | 0.1505 | 1.4939 | 59 |
| HLA-A02:03 | AISSVLNDI | AISSVLNDI | 0.1492 | 1.5057 | 60 |
| HLA-A02:03 | YVTQQLIRA | YVTQQLIRA | 0.1446 | 1.5469 | 61 |
| HLA-A02:03 | QMAYRFNGI | QMAYRFNGI | 0.1436 | 1.5557 | 62 |
| HLA-A02:03 | LLINNATNV | LLIVNNATNV | 0.1423 | 1.5671 | 63 |
| HLA-A02:03 | TLDSKTQSL | TLDSKTQSLL | 0.1415 | 1.5743 | 64 |
| HLA-A02:03 | FQFCNDPFL | FQFCNDPFL | 0.1385 | 1.6011 | 65 |
| HLA-A02:03 | LIANQFNSA | LIANQFNSA | 0.1382 | 1.6036 | 66 |
| HLA-A02:03 | ALGKLQDVV | ALGKLQDVV | 0.138 | 1.6049 | 67 |
| HLA-A02:03 | KLNESLIDL | KNLNESLIDL | 0.1369 | 1.6147 | 68 |
| HLA-A02:03 | KEIDRLNEV | KEIDRLNEV | 0.1283 | 1.6881 | 69 |
| HLA-A02:03 | AQFNGLTVL | AQKFNGLTVL | 0.1209 | 1.7612 | 70 |

| | | | | | |
|---|---|---|---|---|---|
| HLA-A02:03 | RLPQGFSAL | RDLPQGFSAL | 0.1209 | 1.7616 | 71 |
| HLA-A02:03 | SALGKLQDV | SALGKLQDV | 0.1205 | 1.7656 | 72 |
| HLA-A02:03 | TLDSKTQSL | TTLDSKTQSL | 0.1201 | 1.7703 | 73 |
| HLA-A02:03 | AQYTSALLA | AQYTSALLA | 0.1198 | 1.7729 | 74 |
| HLA-A02:03 | IVFPNITNL | IVRFPNITNL | 0.1196 | 1.7749 | 75 |
| HLA-A02:03 | AVRDPTLEI | AVRDPQTLEI | 0.1188 | 1.7834 | 76 |
| HLA-A02:03 | VLNDILSRL | VLNDILSRL | 0.1176 | 1.7953 | 77 |
| HLA-A02:03 | KIQDSLSST | KIQDSLSST | 0.116 | 1.8168 | 78 |
| HLA-A02:03 | KIQDSLSTA | KIQDSLSSTA | 0.1116 | 1.8837 | 79 |
| HLA-A02:03 | YLQPR-TFL | YLQPRTFL | 0.1106 | 1.8977 | 80 |
| HLA-A02:03 | GISGINASV | GDISGINASV | 0.11 | 1.9067 | 81 |
| HLA-A02:03 | QLSSNFGAI | QLSSNFGAI | 0.1063 | 1.954 | 82 |
| HLA-A02:03 | ALIPFAMQM | ALQIPFAMQM | 0.1041 | 1.9827 | 83 |
| HLA-A02:03 | LLH-APATV | LLHAPATV | 0.1031 | 1.9958 | 84 |

**Table S5. Strong HLA A*02:01 or A*02:03 binding epitopes in SARS-CoV-2 spike (predicted using NetMHCpan v4.1)**

| Spike Epitope | A*02:01 (% Rank) | A*02:03 (% Rank) | Notes |
|---|---|---|---|
| VLNDILSRL | 0.028 | 0.0058 | |
| LLFNKVTLA | 0.1053 | 0.0391 | |
| YLQPRTFLL | 0.0129 | 0.0406 | |
| HLMSFPQSA | 0.1085 | 0.0611 | |
| ALNTLVKQL | 0.2258 | 0.0626 | |
| VVFLHVTYV | 0.1528 | 0.084 | |
| RLNEVAKNL | 0.2303 | 0.0907 | |
| RLQSLQTYV | 0.0622 | 0.0939 | |
| FIAGLIAIV | 0.2409 | 0.0997 | |
| TLDSKTQSL | 0.0414 | 0.103 | |
| SVLNDILSRL | 1.3286 | 0.1387 | Enhanced binding for A*02:03 |
| KLNDLCFTNV | 0.3539 | 0.156 | |
| VLYENQKLI | 0.3975 | 0.1785 | |
| RLITGRLQSL | 2 | 0.2509 | Enhanced binding for A*02:03 |
| KIADYNYKL | 0.0671 | 0.2647 | |
| SIIAYTMSL | 0.2998 | 0.27 | |
| KLPDDFTGCV | 0.3198 | 0.2722 | |
| RVYSTGSNV | 1.942 | 0.3381 | Enhanced binding for A*02:03 |
| SLSSTASAL | 0.9509 | 0.3881 | Enhanced binding for A*02:03 |
| YHLMSFPQSA | 0.4737 | 0.3955 | |
| KLQDVVNQNA | 0.511 | 0.4003 | Enhanced binding for A*02:03 |
| TVYDPLQPEL | 2 | 0.4133 | Enhanced binding for A*02:03 |
| KIYSKHTPI | 1.3468 | 0.4213 | Enhanced binding for A*02:03 |
| FTISVTTEI | 0.575 | 0.4596 | Enhanced binding for A*02:03 |
| FELLHAPATV | 0.3775 | 0.4617 | |
| NLNESLIDL | 0.2624 | 0.4634 | |
| LLALHRSYL | 1.942 | 0.4767 | Enhanced binding for A*02:03 |
| RLDKVEAEV | 0.0899 | 0.4814 | |
| VLSFELLHA | 0.627 | 0.4893 | Enhanced binding for A*02:03 |
| GLTVLPPLL | 0.2593 | 0.6876 | Enhanced binding for A*02:01 |
